# Developing an automatic system for classifying chatter about health services from Twitter: A case study for Medicaid

**DOI:** 10.1101/2020.06.12.20129593

**Authors:** Yuan-Chi Yang, Mohammed Ali Al-Garadi, Whitney Hogg-Bremer, Jane M. Zhu, David Grande, Abeed Sarker

**Affiliations:** Department of Biomedical Informatics, School of Medicine, Emory University, Atlanta, GA; Division of General Internal Medicine and Geriatrics, Oregon Health & Science University, Portland, OR; Division of General Internal Medicine, Perelman School of Medicine, University of Pennsylvania, Philadelphia, PA; Department of Biomedical Engineering, Georgia Institute of Technology and Emory University, Atlanta, GA

**Keywords:** Natural Language Processing, Machine Learning, Twitter, Social Media, Medicaid, Consumer Feedback

## Abstract

**Background:** The wide adoption of social media in daily life renders it a rich and effective resource for conducting close-to-real-time assessments of consumers’ perceptions about health services. This is, however, challenging due to the vast amount of data and the diverse content in the social media chatter.

**Objectives:** To develop and evaluate an automatic system, involving natural language processing and machine learning, for automatically characterizing user-posted Twitter data about healer services, using Medicaid, the single largest insurance in the United States, as an example.

**Methods:** We collected data from Twitter in two ways: (i) via the public streaming API using Medicaid-related keywords (Corpus-1), and (ii) by using the website’s search option for tweets mentioning the agency-specific handles (Corpus-2). We manually labeled a sample of tweets into five pre-determined categories or *other*, and artificially increased the number of training posts from specific low-frequency categories. Using the manually-labeled data, we trained and evaluated several supervised learning algorithms, including Support Vector Machine, Random Forest (RF), Naïve Bayes, shallow Neural Network (NN), k-Nearest Neighbor, Bi-Directional Long Short-Term Memory, and Bidirectional Encoder Representations from Transformers (BERT). We then applied the best-performing classifier to the collected tweets for post-classification analyses assessing the utility of our methods.

**Results:** We manually annotated 11,379 (Corpus-1: 9,179; Corpus-2: 2,200) tweets, using 7,930 (69.7%) for training and 1,449 (12.7%) for validation and 2,000 (17.6%) for test. A BERT-based classifier obtained the highest accuracies (81.7%, Corpus-1; 80.7%, Corpus-2) and F_1_-score on Consumer Feedback (0.58, Corpus-1; 0.90, Corpus-2), outperforming the second-best classifiers in accuracies (74.6%, RF on Corpus-1; 69.4%, RF on Corpus-2) and F_1_-score on Consumer Feedback (0.44, NN on Corpus-1; 0.82, RF on Corpus-2). Post-classification analyses revealed differing inter-corpora distributions of tweet categories, with political (64%) and consumer-feedback (55%) tweets being the most frequent for Corpus-1 and -2, respectively.

**Conclusions:** The broad and variable content of Medicaid-related tweets necessitates automatic categorization to identify topic-relevant posts. Our proposed system presents a feasible solution for automatic categorization, and can be deployed/generalized for health service programs other than Medicaid. Annotated data and methods are available for future studies (https://yyang60@bitbucket.org/sarkerlab/medicaid-classification-script-and-data-for-public).

## Introduction

Consumers’ perspectives and feedback are crucial for improving products or services. Over the last two decades, widespread adoption and use of the Internet has led to its utilization as a major platform for collecting targeted consumer feedback. Businesses often allow consumers to rate specific products and/or services, and also provide detailed comments or reviews, and this has become a key feature of e-commerce platforms. For example, consumer-generated reviews and ratings of products play an important role in differentiation on Amazon, which currently has a global presence.^1,2^ There are also companies, such as Yelp, that focus specifically on enabling crowdsourcing consumer feedback.^3-6^ Similarly, as social media has become the primary platform of communication for many people, many companies have started maintaining and communicating via social media accounts, often enabling direct communications, both private and public, with consumers. Not only do consumers provide comments or seek assistance through those social media accounts, but they also often engage in discussions about products or services within their own social networks. Consequently, such consumer-generated chatter is often utilized to assess their perceptions of specific topics, which may range from products or services to social programs, legislations and politicians.

Social media is a rich resource for obtaining perspectives on public health, since it enables the collection of large amounts of data directly and in real-time. It is commonly used for sentiment analysis—a field of study that analyzes opinions, sentiments, attitudes and emotions from written language. Sentiment analysis research involving social media data has covered a wide range of topics, events, individuals, issues, services, products, and organizations.^7,8^ The use of social media has not, however, been limited to sentiment analysis in open domains. Over recent years, research within the broader medical domain have embraced social media, and it is currently being utilized for conducting real-time public health surveillance, including for topics such as influenza surveillance, pharmacovigilance and toxicovigilance.^9-11^ Meanwhile, similar to corporate businesses in the United States (US), health service providers such as local health departments and hospitals have also started adopting social media specifically as a consumer-facing communication channel.^12,13^ Prior studies in this space have investigated how the social media data linked to such health services accounts reflects the consumers’ perspectives about them. The simplest studies have focused on utilizing structured or numeric information, such as likes or ratings, associated with the accounts belonging to hospitals or nursing homes, and these metrics had been compared against traditional quality reports and ratings.^14-16^ Building on the advances in open-domain natural language processing (NLP), some studies within the broader health domain have attempted to use unstructured data, including postings related to patient experiences about hospitals, to infer consumer sentiments^17,18^ or extract topics that summarize content.^19^

Extracting knowledge from social media data is notoriously difficult for NLP methods due to factors such as the presence of misspellings, colloquial expressions, lack of context, and noise. These problems are exacerbated for health-related data due to the complexities of domain-specific terminologies, the lack of expert knowledge among common social media users, and the uniqueness of health-related topics. Consequently, there is considerably less research exploiting social media free text data for health-related tasks. Past studies closely related to ours have focused on analyzing sentiments towards attributes of health insurance plans^20^ and social media users’ responses to public announcements about health policies.^21^ However, to the best of our knowledge, there have been no close-to-real-time automatic system that provides comprehensive data collection and analysis on social media chatter about health services and insurance coverage provided by large public insurers such as Medicaid and Medicare. This is perhaps because social media chatter associated with these topics cover more diverse perspectives, which increase the complexities posed by them to NLP methods. For example, for a medical entity such as a hospital, it is easy, from a relative perspective, to characterize the chatter in terms of user sentiment (*e.g*., how the user feels about the services provided by the hospital). However, chatter associated with an entity like Medicaid contains discussions about politics and legislations, academic research, statistics and factual information, consumer feedback, and so on. Chatter related to politics will be different, in terms of content, compared to chatter related to consumer feedback. In fact, sentiment may also have different meanings for these two broad categories of chatter—negative sentiment in political chatter may represent a user’s emotions associated with a political decision about the health service (*e.g*., changes in policies related to insurance coverage or covered benefits within Medicare or Medicaid), rather than the service itself.

Due to these additional complexities, there is a need to identify the broad categories of information in such social media chatter before they can be used for targeted analyses, such as sentiment analyses. These complexities, combined with the promise of social media data and the lack of past research in this specialized area, served as the primary motivation for the study described in this paper. We chose Medicaid as our target health service because it is the single largest public insurance program in the country^22^ and contains large volumes of related chatter on social media.

The specific objectives of this paper are to:

- Assess if a social media platform, specifically Twitter, contains sufficient volumes of chatter about health services so that it can be used to conduct largescale analyses, using Medicaid as our target service.
- Develop and discuss a data-centric system, involving NLP and machine learning, for automatically collecting, categorizing and analyzing Twitter chatter associated with Medicaid, shown in Figure 1.
- Describe the manual annotation of a Twitter-Medicaid dataset, and its composition.
- Describe supervised classification strategies for automatically classifying Medicaid-related tweets into broad categories, and evaluating the performances of several machine learning models, with particular emphasis on tweets that potentially represent consumer feedback.
- Conduct post-classification content analyses to verify the potential utility of our data-centric system.

**Figure 1.**
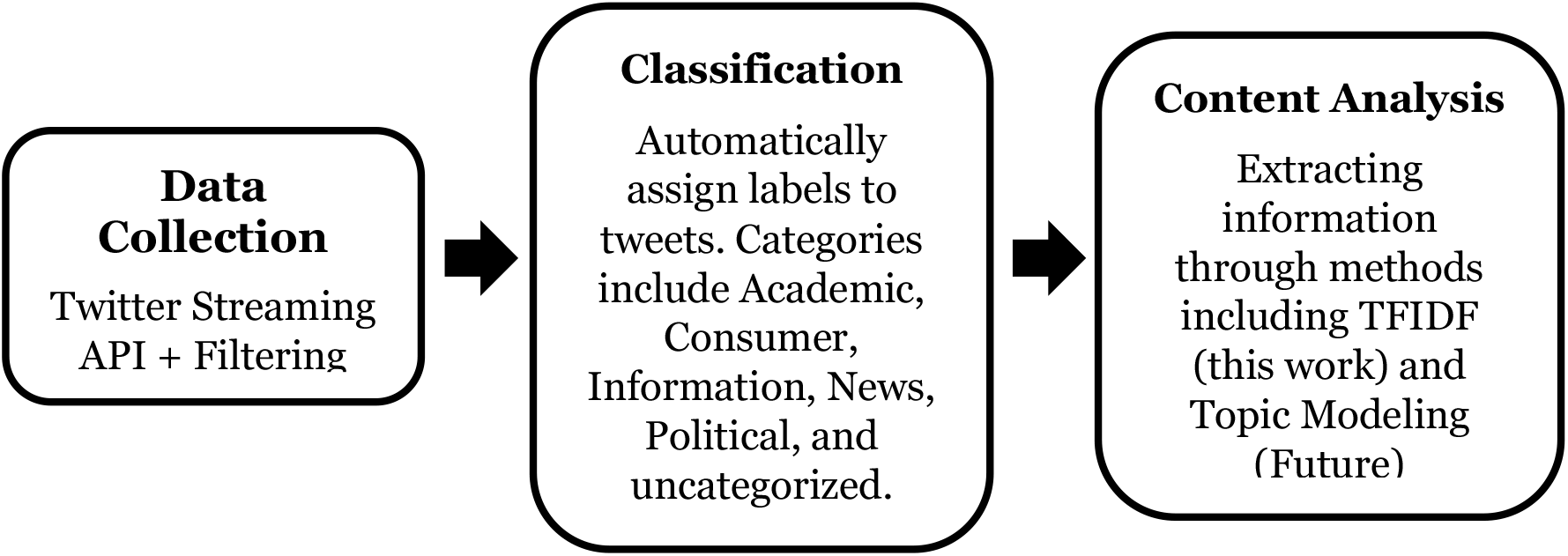
The NLP System for automatic data collection, classification, and content analysis of the Medicaid chatter on Twitter

The main contributions of this paper are as follows:

- We present the methods and results of collecting Medicaid-related Twitter data, analyzing a sample of the data manually, and developing an annotation guideline suitable for preparing a large dataset for training classification algorithms.
- We present details of automatic supervised classification experiments, including methods, results and evaluations, and provide suggestions about how to further improve the performance.
- We discuss the post-classification analyses of the collected data, including data distribution and content analyses.
- We make the NLP and machine learning scripts in this work publicly available, along with the labeled training dataset and a larger set of unlabeled Medicaid-related data.

## Methods

### Data Collection

To develop our models for the analyses of data related to Medicaid from Twitter, we collected two sets of publicly available data from the network, which we labeled as Corpus-1 and Corpus-2. Corpus-1 contains tweets mentioning the term ‘*medicaid*,’ or Medicaid agency (MA) and managed care organization (MCO, an organization that provides Medicaid-related health services under contracts from the agency) names that are branded and thus easily distinguishable on Twitter (e.g., *Medi-cal*: California’s Medicaid program; and *TennCare*, Tennessee’s Medicaid program). These tweets were collected via Twitter’s public streaming API (application programming interface) from May 1, 2018 to October 31, 2019, limited to only English tweets. As it is reported that misspellings appear frequently on social media platforms,^23^ particularly Twitter, we used an automatic spelling variant generator to generate common misspellings for ‘*medicaid*’ and used them to capture tweets referring *‘medicaid’* as one of the misspellings.^24^ This can increase retrieval rate and increase the volume of the streaming data. The full list of keywords, including the misspellings, are shown in Table 1 in the Supplementary Material. We then identified and removed tweets whose contents were not directly related to Medicaid and repeated/duplicated contents/tweets (*e.g*., fund-raising or political campaign). To focus on tweets expressing personal opinion, we also removed retweets, deemed as duplicates of the original tweets. The final dataset consisted of 628,411 tweets for Corpus-1. While most of the chatter regarding Medicaid posted by consumers only included the term ‘*medicaid*’ (or its variants), some directly tagged or mentioned relevant Twitter handles associated with MAs or the MCOs (e.g., ‘@organization_name’). Corpus-2 is composed of such tweets, and the MA and MCO Twitter handles were identified in a previous study.^25^ The full list of the handles used in data collection is presented as Table 2 in the Supplementary Material. These tweets were retrieved by targeted searching ‘(*e.g*., *to:organization_name*)’ on Twitter. These tweets were posted between December 12, 2008 and the time of search (January 9, 2020). We filtered the tweets using the same approaches that were used for Corpus-1. In total, there are 27,337 tweets in the corpus. Additional notes about our data is provided in the Supplementary Materials.

**Table 1.** Distribution (counts and percentages) for annotated data in the first round of annotations (row 2 & 3), and the final data sets (Corpus-1 for row 4 and 6; Corpus-2 for row 5 and 7)

| Category | Academic | Consumer | Information | News | Political | Other | Total |
| --- | --- | --- | --- | --- | --- | --- | --- |
| Training set (1<sup>st</sup> round) | 61 (1.1%) | 158 (2.7%) | 198 (3.4%) | 1288 (22.2%) | 3613 (62.3%) | 477 (8.2%) | 5795 |
| Validation set (1<sup>st</sup> round) | 35 (2.4%) | 37 (2.6%) | 49 (3.4%) | 317 (21.9%) | 897 (61.9%) | 114 (7.9%) | 1449 |
| Training set (Corpus-1) | 83 (1.2%) | 355 (5.3%) | 429 (6.4%) | 1299 (19.3%) | 3710 (55.1%) | 854 (12.7%) | 6730 |
| Training set (Corpus-2) | 9 (0.8%) | 709 (59.1%) | 94 (7.8%) | 40 (3.3%) | 10 (0.8%) | 338 (28.2%) | 1200 |
| Test set (Corpus-1) | 20 (2%) | 46 (4.6%) | 49 (4.9%) | 199 (19.9%) | 603 (60.3%) | 83 (8.3%) | 1000 |
| Test set (Corpus-2) | 6 (0.6%) | 579 (57.9%) | 80 (8%) | 21 (2.1%) | 6 (0.6%) | 308 (30.8 %) | 1000 |
| Total | 153 | 1726 | 701 | 1876 | 5226 | 1697 | 11379 |

**Table 2.**
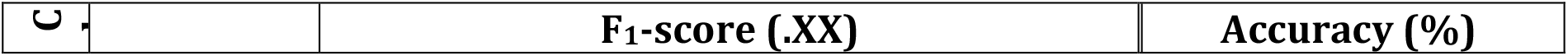
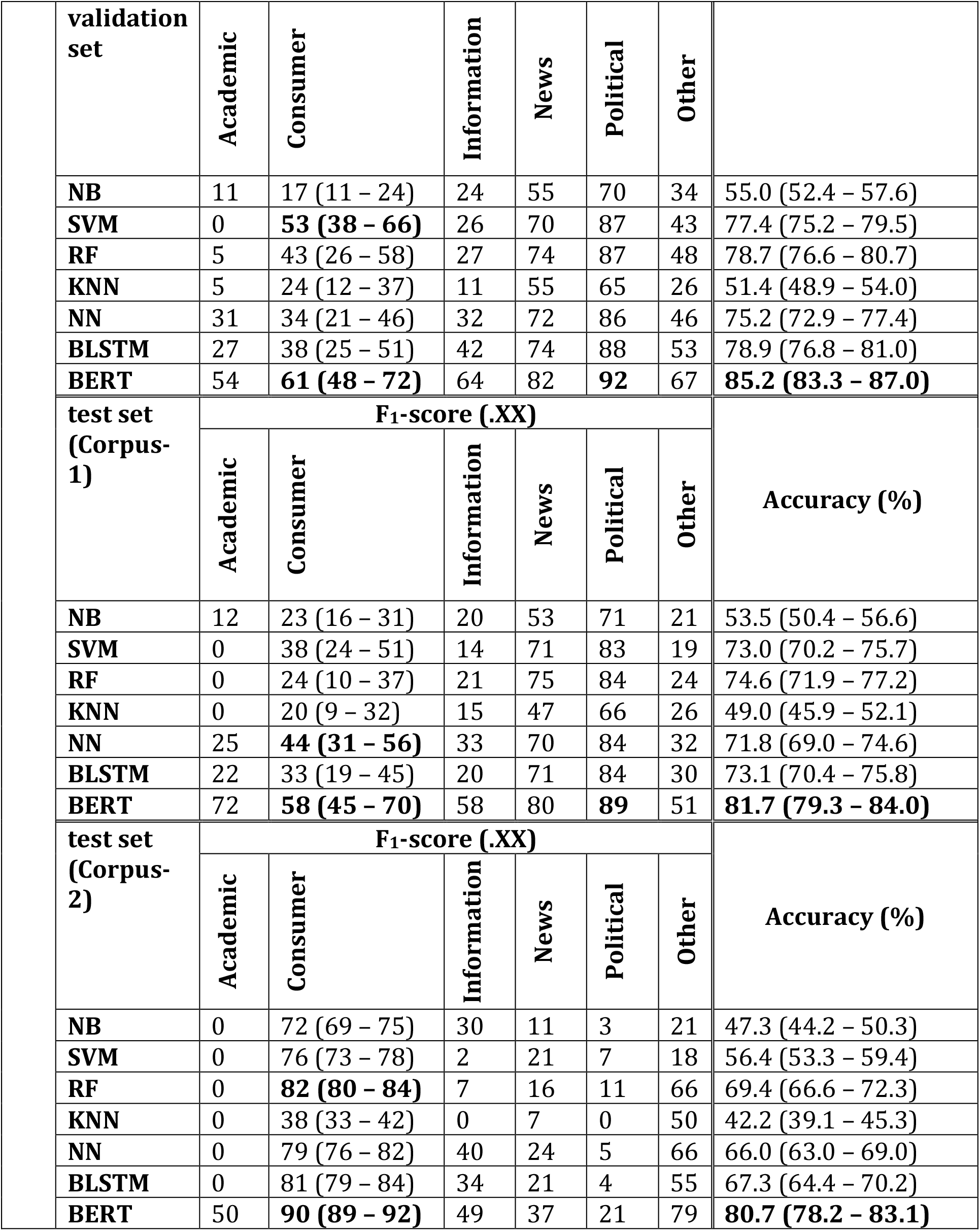
Classification performances of the classifiers on the test sets of Corpus-1 & 2. The 95% confidence intervals are given in parenthesis.

### Tweet Contents and Manual Annotations

To better understand the contents of the tweets posted by users, and to develop methods to automatically characterize the posts, we first performed manual inspections of the contents of the posts and identified commonly occurring themes. We used the grounded theory approach to conduct a thorough analysis^26^—we analyzed a random sample of tweets to identify recurring topics, and then grouped the topics into broader categories/themes. The analysis was conducted by multiple authors of this paper and the topics discovered initially were discussed. The discovered topics were either merged into broader themes (*e.g*., combining *information* and *outreach*), discarded from our consideration (*e.g*., for topics that were observed rarely or once-only), or split into multiple themes (*e.g*., splitting of *information* tweets into *academic, information/outreach*, and *news*). We eventually settled for 5 broad categories, which were decided upon following discussions and finalized by the domain-expert authors of this paper (JMZ, DG): (i) academic, (ii) consumer feedback, (iii) information/outreach, (iv) news, and (v) political opinion/advocacy. Tweets that could not be categorized as any of these were labeled as *other*. Descriptions of these classes are provided below:

- **Academic (Academic):** Tweets related to research about Medicaid. These include tweets by persons or organizations with academic affiliations or think tanks that express the perspective from the affiliated organizations, or any tweet relating to education, scholarship, and thought including (links to) journal publications and reports.
- **Consumer Feedback (Consumer):** Tweets related to consumers’ experiences or questions related to Medicaid services, coverage, benefits, or health issues. The tweets are typically from Medicaid consumers or family members of consumers, and can also include discussions with others.
- **Information/Outreach (Information):** Tweets directed at consumers and beneficiaries of Medicaid to convey information including agency services, programs, events, enrollment, eligibility criteria, etc. The tweets containing information about general health or public health reminders are also included.
- **News (News):** News and announcements–including any tweets from a news agency/organization. The tweets that explicitly express political opinions and the tweets from Medicaid agencies/plans are excluded.
- **Political opinion / advocacy (Political):** Comments, personal opinions, and feedback about politics related to Medicaid.
- **Other (Other):** Tweets that are not relevant, typically the noise that isn’t captured by the initial screening.

Following the establishment of the desired categories and the development of annotation guidelines by JMZ, two trained annotators performed a first round of annotations (for the data in Corpus-1) in multiple iterations, developed annotation guidelines, and resolved ambiguities via discussion. Following the completion of this round of annotations, the annotation disagreements were resolved by AS and WH. We found the class distribution to be very imbalanced, with most of the tweets annotated as *News, Political*, and *Other*, while only a small portion were in *Academic, Consumer*, and *Information* (Table 1). To understand how this imbalanced distribution affected the classifier performances on the smaller classes, particularly the *c* class, we performed preliminary automatic classification experiments using three classifiers: Naïve Bayes (NB), Support Vector Machine (SVM) and Random Forest (RF). We split the data into training (80%) and validation (20%) sets, and found the best performance on *Consumer Feedback* to be low for all the classifiers, with best F1-score = 0.3 (SVM). Tweets belonging to the *Consumer Feedback* class was of particular importance to our overarching project objectives, so we devised two strategies for improving performance for this class—the first involving additional annotations of targeted tweets from the same dataset, and the second focusing on collecting an additional dataset (Corpus-2, as described earlier).

For the first strategy, we conducted another round of annotation of tweets from Corpus-1 to increase the number of tweets for the *c* class. Due to the very low numbers of *c* class tweets in the original dataset, we realized that it would not be feasible to annotate sufficient numbers of these tweets by drawing random samples because of budgetary and other constraints. Therefore, rather than randomly drawing tweets for the next round of annotations, which would again lead to finding a small number of tweets belonging to *Consumer Feedback* category, we attempted to artificially increase the number of tweets for this category. We achieved this by running our above-described *weak* classifier on a larger set of unlabeled tweets and only picking tweets classified as *Consumer Feedback* by the SVM classifier. This significantly increased the number of *Consumer Feedback* tweets in the data to be annotated. The new set of annotated data were then added into the training set, and the data distribution is presented in Table 1.

We followed the similar annotation strategy for Corpus-2 (i.e., annotating tweets classified as Consumer by the classifier trained on the previously annotated data), but this time we also annotate equal amount of non-Consumer tweets. This is because Corpus-2 is rich in Consumer tweet and we would also like to include tweets in other categories to improve the performance. An outline of the overall annotation process is presented in Figure 2. While we tried to decrease the class imbalance in the training sets of the two corpora, to ensure that our evaluations represented the classifier performances on real-world distributions of the data, we did not artificially balance the validation set. We also annotated the test set randomly generated from the two corpora, 1,000 tweets each, so they would reflect the data composition of the original corpora, allowing us to evaluate how the classifier would perform when deployed, especially when taking streaming data.

**Figure 2.**
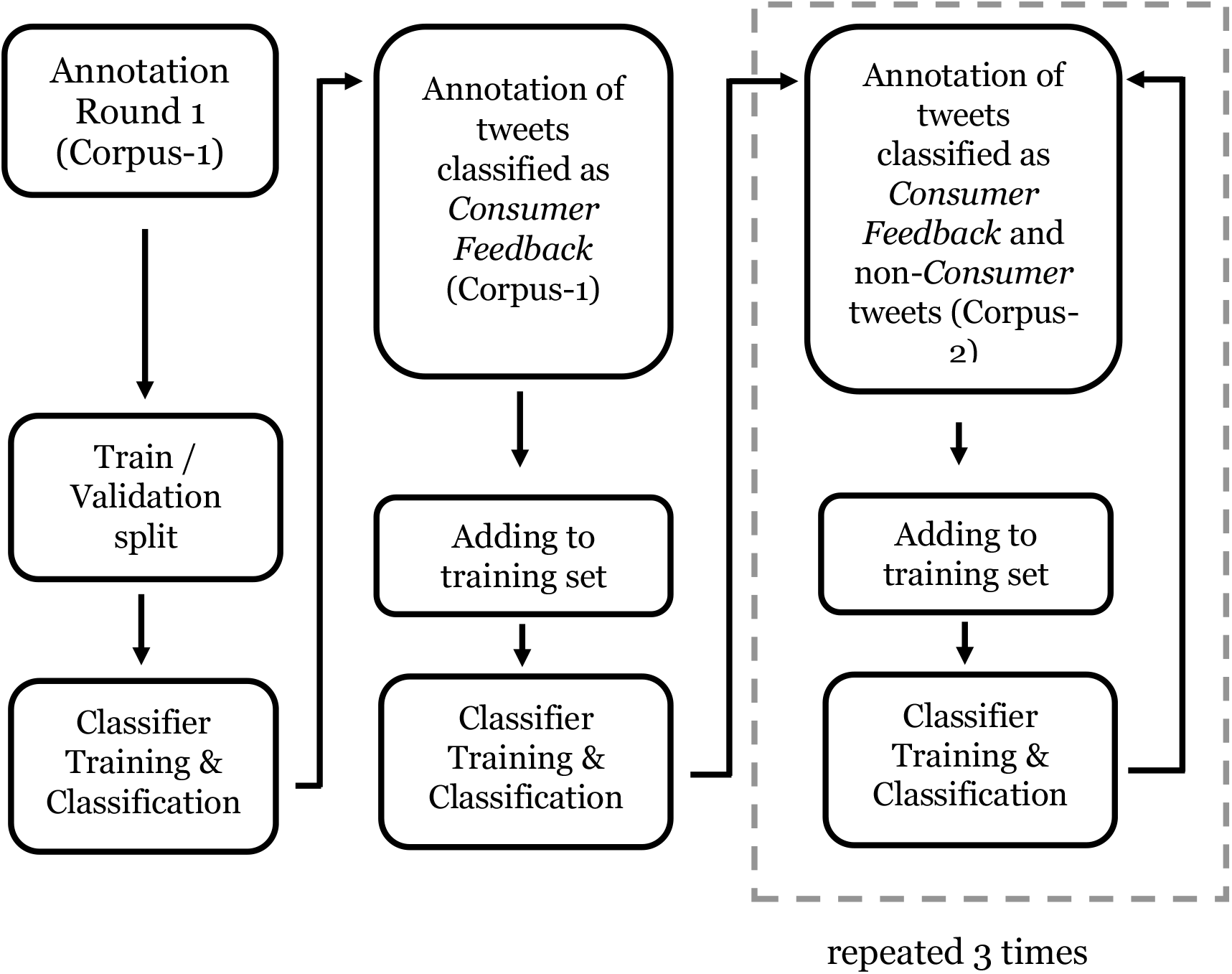
Flow chart for the entire annotation process involving multiple rounds

### Classification

We experimented with five traditional classification algorithms, including Gaussian NB,^27,28^ SVM,^29,30^ RF,^31^ k-Nearest Neighbor (KNN),^28^ and shallow Neural Networks (NN), and two advanced classification algorithms, Bi-directional Long Short-Term memory (BLSTM)^32,33^ and Bidirectional Encoder Representations from Transformers (BERT)^34,35^. Although the origin and distributions of tweets in the two corpora were different, we decided to combine them since our past research suggests that multi-corpus training, or distant supervision, leads to performance improvements for social media text classification.^36^ The feature extraction and classification training for traditional classifier is done using the “Scikit-learn” package in python,^37^ the BLSTM classification is implemented using package “Keras” in python,^38^ and the BERT classification is implemented using package “simpletransformers” which is based on the package “transformers.”^39^ The performance on the validation set and the test set from Corpus-1 and Corpus-2 are shown in Table *2*.

The tweets were pre-processed by lowercasing and anonymizing URLs and user names. For the traditional classifiers, the non-English characters were further removed, and each word was stemmed by the Porter stemmer. The features were the un-normalized counts of the 3000 most frequent n-grams (contiguous sequences of words with n ranging from 1 to 3, with 1380 unigrams, 1296 bigrams, and 324 trigrams). We also introduced a “word cluster” feature, which are clusters or generalized representations of semantically similar words/phrases learned from Twitter chatter^40,41^. The word clusters were represented as bag-of-word vectors, and the feature space consisted of 972 word clusters. We used the Twitter word clusters, “*50mpaths2*,” provided by Owoputi et al (2012).^42^ For the advanced classifiers, each word or character sequence was replaced with a dense vector, and the vectors were then fed into the relevant algorithms for training.

We performed hyper-parameter tuning using the validation set for improving the classification task on the imbalanced dataset. Specifically, we focused on improving the F_1_-score for *Consumer Feedback*. For traditional classifiers, we optimized number of nearest neighbors for KNN, the number of estimators (trees) for RF, and the weights for SVM. We have also experimented with over-sampling using Synthetic Minority Over-sampling Technique (SMOTE) but performance was not improved (provided in the Table 3 in the supplementary materials). The optimal hyperparameters for the traditional classifiers are given in the Table 4 in the supplementary materials. We used Twitter GloVe word embeddings for the BLSTM^43^ classifier, where each word is converted to 200-dimensional vector. BLSTM was then trained with 40 epochs and dropout regularization and the best model was selected through accuracy on validation data. We chose RoBERTa-large for BERT algorithms,^35^ trained with 3 epochs. The technical details are provided in the Table 4 in the Supplementary Materials.

**Table 3.** BERT Classifier’s confusion matrix on test set.

| test set (Corpus-1) |  | Predicted Value |  |  |  |  |  |
| --- | --- | --- | --- | --- | --- | --- | --- |
|  |  | Academic | Consumer | Information | News | Political | Other |
| True Value | Academic | 13 | 0 | 1 | 4 | 2 | 0 |
|  | Consumer | 0 | 26 | 0 | 0 | 18 | 2 |
|  | Information | 0 | 0 | 27 | 9 | 10 | 3 |
|  | News | 1 | 0 | 9 | 169 | 17 | 3 |
|  | Political | 2 | 5 | 3 | 39 | 549 | 5 |
|  | Other | 0 | 12 | 4 | 4 | 30 | 33 |
| test set (Corpus-2) |  | Predicted Value |  |  |  |  |  |
|  |  | Academic | Consumer | Information | News | Political | Other |
| True Value | Academic | 3 | 0 | 0 | 1 | 2 | 0 |
|  | Consumer | 1 | 512 | 1 | 2 | 26 | 37 |
|  | Information | 1 | 5 | 33 | 15 | 1 | 25 |
|  | News | 0 | 0 | 5 | 11 | 3 | 2 |
|  | Political | 0 | 0 | 0 | 1 | 5 | 0 |
|  | Other | 1 | 36 | 15 | 7 | 6 | 243 |

### Post-classification Analyses

To assess the utility of out classification approaches and gain understanding of the data, we used the best-performing classifier (the classifier based on BERT) to label all collected unlabeled data and compute the data distribution. We then performed content analysis using the term-frequency-inverse-document-frequency method (TFIDF),^44^ focusing on the tweets in Corpus-1 that contained the term ‘*medicaid*’ and its misspellings. Our intent was to qualitatively assess that the classifier was capable of distinguishing tweets based on contents that are manually verifiable. For all content analyses, the text was first pre-processed by lowercasing, removing URLs, user names, non-English characters, stopwords, and any word with less than four characters.

## Results

### Annotation and Class Distributions in test sets

We annotated a total of 9,179 tweets from Corpus-1 and 2,200 tweets from Corpus-2. We obtained substantial inter-annotator agreement (Cohen’s κ = 0.734) ^45,46^ over 892 double-annotated tweets. The test data sets were randomly selected from the corpora and, therefore, can be considered a sample of the collected data. For Corpus-1, the test data contained 1,000 tweets, among which the *political discussion* (class p) was the dominant class (60.3%) followed by *news* (class n; 19.9%), while *consumer feedbacks* (class c) made up less than 5% of the tweets. In contrast, consumer feedback comprised about 58% in Corpus-2 and roughly 31% of the tweets could not be categorized, most of which were part of conversations and could not be understood without full context.

### Classification Results

The F1-scores for each class and the accuracies of the classifiers on the validation set and the test sets are presented in Table *2*, including confidence intervals estimated using bootstrapping, while the precisions and the recalls are given in Table 5 in the Supplementary Materials due to space limit. For the validation set and the test set from Corpus-1, the classifiers showed high performance for class p, but relatively lower for class c. This was expected based on the large imbalance described earlier. Among all the traditional classifiers experimented, SVM performed the best on the validation set, with F_1_-score of 0.53 on the Consumer Feedback. However, the F_1_-score on the Consumer Feedback on the test set from Corpus-1 is only 0.38. In contrast, we found that the BERT classifier has the highest F_1_-scores on Consumer Feedback for both the validation set (0.61) and the test set from Corpus-1 (0.58).

For the test set from Corpus-2, most of the classifiers performed well on the *Consumer Feedback*. Among the traditional classifier, RF performed the best, with F_1_-score of 0.82 on *Consumer Feedback*. On the other hand, BERT still performed the best, with Consumer Feedback F_1_-score of 0.90.

Since BERT classifier performed the best in the accuracy and the Consumer Feedback F_1_-score on the validation set as well as the two test sets, we used the BERT classification for post-classification analysis.

### Error analysis

We conducted a brief analysis of the errors made by the BERT-based classifier. We first calculated the confusion matrix on both test sets (Table 3). In Table 4, we provided examples for the most frequent classification errors, omitting the unnecessary details. For Corpus-1, we highlighted that the classifier frequently misclassified *Political* tweets as *News*, or *Consumer Feedback*, and vice versa. This is not surprising because users sometimes commented and discussed politics with personal experience and some news content was related to opinions about the policy. We also highlighted that the uncategorized tweets, whose content is often not directly related to Medicaid or lack of information, are frequently misclassified as *Consumer Feedback* or *Political*. The confusion between *Consumer Feedback*, and *Political* or uncategorized tweets, along with the low volume of *Consumer Feedback*, contributes to the low performance on the *Consumer Feedback*. We also observed that some *News* tweets were confused with the *Information* tweets because information is frequently spreading as news or blog articles. For Corpus-2, the dominating classes were *Consumer Feedback* and uncategorized tweets, and they were most frequently misclassified as each other. We suspect they were misclassified because tweets sometimes lacked context, making their meanings ambiguous and hard for the machine to understand. For example, the tweet ‘*<organization_name> poorly worded,’* though ambiguous, might be understood as that some document for customer or the customer service representative’s expression was poorly worded, and thus, we categorized it as *Consumer Feedback*. However, the machine learning algorithms were not capable of deciphering such implicit contexts—that ‘poorly worded’ is usually associated with a feedback, and, in tweets directed to the agency’s handle, it is likely related to customer service. Similarly, the tweet *‘<organization_name> My pleasure!*,’ may belongs to a conversation between a customer and a representative but the lack of information renders it to the *Other* class. However, the machine learning were not able to capture this understanding.

**Table 4.** Examples of Misclassified tweets by BERT Classifier on Corpus-1 and Corpus-2

|  |  |  |  |
| --- | --- | --- | --- |
| test set (Corpus -1) | Tweets | True class<br>(prediction) | Comments |
|  | I need this government shutdown to end because no one is going to call me to set up my Medicaid while it's shutdown | *political*<br>(*consumer*) | Discussion about politics with personal experience |
|  | 'This is just cruelty and exclusion': Amid Trump's attack on poor, one million fewer kids receiving Medicaid and CHIP – Raw Story <URL> | *political*<br>(*news*) | Opinion on Medicaid policy presented as a news title |
|  | <USERNAME> So do I! But I totally understand why some people really hate it. And yes... lack of Medicaid providers is a problem everywhere (I do accept it, but only have a mobile practice). Maybe contact your local health department and ask! | *consumer*<br>(*political*) | Customer's discussion about Medicaid services. It might be misclassified due to similarity to the Medicaid political discussion |
|  | Thanks to <USERNAME> for this story about the bill ... Ohio leaving some military families with special needs children waiting for answers <URL> | *news*<br>(*political*) | News about Medicaid policy reformation bill |
|  | States that been successful in lowering substance use disorder rates have increased access to medicaid & private insurance, and to MAT and naloxone. Thank you, NYT Editorial Board @NYTOpinion. <URL> | *news*<br>(*information*) | News about information related to Medicaid |
|  | 3 Ways to increase Missouri Medicaid EMOMED Reimbursement <URL> | *Information*<br>(*news*) | Information for Medicaid beneficiaries, presented as a blog article |
|  | The Medicaid office I'm going to tomorrow opens at 7:30 am. I won't be there that early, but ugh. | *other*<br>(*consumer*) | Uncategorized because it is not about experience or question, but content indicates that user to be a customer. |
|  | <USERNAME> I hope someone will ask him " What's the difference between Medicaid and Medicare?" | *other*<br>(*political*) | Uncategorized due to lack of related content, but similar to political discussion |
| test set | Tweets | True class<br>(prediction) | Comments |
|  | <organization_name> poorly worded | *consumer*<br>(*other*) | Most likely comment that customer service but hard to pick up by algorithm. |
|  | <organization_name> My pleasure! | *other*<br>(*consumer*) | Classified as others because of lack of information,' but the algorithm might recognized that it could be a conversation between a customer and a customer representative. |

### Post-classification analyses—data distribution

We applied the best-performing classifier (BERT) to label both corpora. The class distribution obtained is shown in Figure 3. We found that the majority of tweets in Corpus-1 were news (class *n*, 23%) and political discussion (class *p*, 64%), while consumer feedbacks (class *c*) only made up 5%, consistent with the data distribution of the test set of Corpus-1. The data distribution indicates that this corpus suits to analysis of chatters regarding to political discussion or news. For Corpus-2, the majority of the tweets were labeled as consumer feedback (class *c*, 55%) and uncategorized (class *o*, 31%), also consistent with the data distribution in the test set.

**Figure 3.**
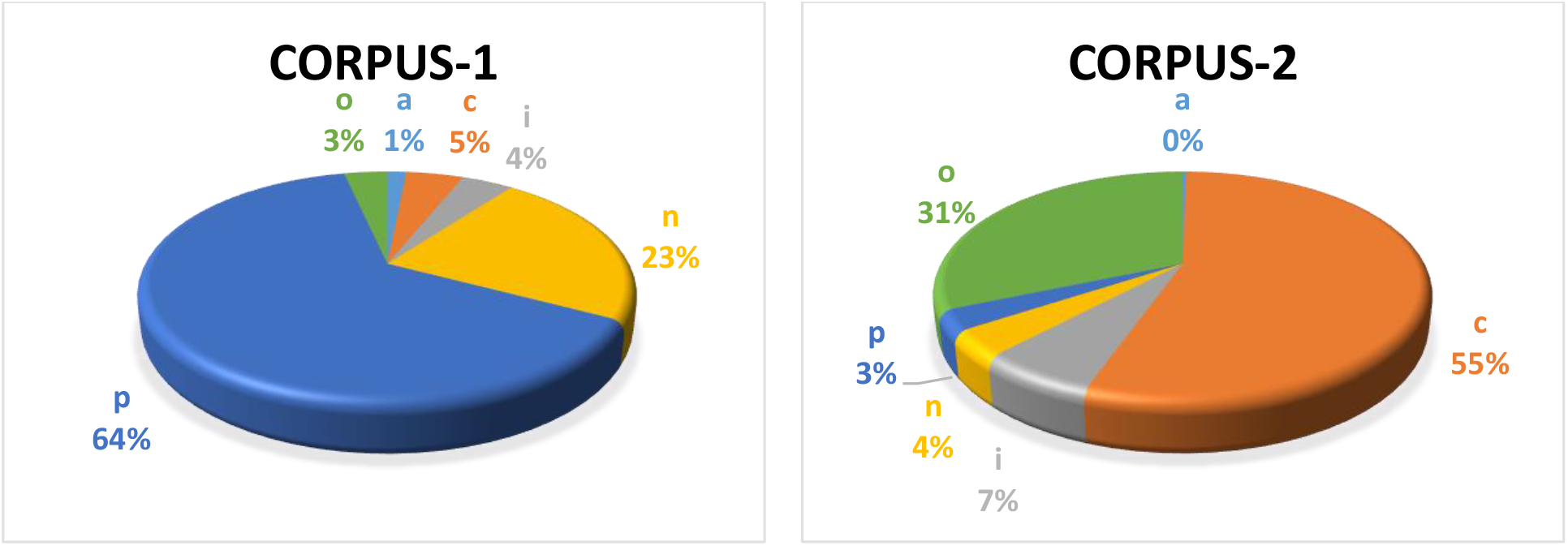
Post-classification class distributions among to two corpora, as per the automatically classified tweets.

### Post-classification Analyses: content of each class in Corpus-1

We now briefly summarize the findings from content analyses on the tweets in Corpus-1 that contain the terms associated with ‘*medicaid*,’ in order to understand, from a high-level perspective, the contents within each category. The 10 highest-ranking bigrams and trigrams detected by TFIDF method are listed in Table 6 in the Supplementary Materials.^44^ Not surprisingly, the Academic tweets (class *a*) are dominated by terms starting with *‘study …’* and terms indicating research finding. Similarly, the *i* class contain terms related to *‘service,’ ‘care,’* … etc, consistent with information outreach. For the *n* class, we found that many tweets were about news on medicaid work requirements in Kentucky and Arkansas (blocked by federal judge on March 27, 2019). In addition, *‘social security’* and *‘Trump …’* are also highly-ranked among the *n* and *p* classes. For the tweets belonging to the *c* class, some of the high-ranking terms were shared with other classes (e.g. *‘… insurance,’ ‘social security,’* or *‘… care’*) while some were specific to this class (*‘make much’* or *‘doesn cover’*) and potentially indicated comments about Medicaid income cap and coverage.

We did not know the compositions of the two datasets we had collected *a priori*. Thus, the results of our classification experiments provided us very important knowledge about which type of Twitter data to use when conducting targeted studies about Health services in general, or Medicaid here. For example, when studying consumer feedback, it is best to use data from Corpus-2 (*i.e*., tweets containing Twitter handles of the MA or MCO); for studying public perceptions of political decisions, Corpus-1 would be more useful. Detailed content analyses of the tweets in each category, such as their temporal and geolocation-specific distributions are likely to reveal more relevant information. However, such analyses is outside the scope of the current study, and we plan to build on the NLP system described in this paper to conduct more thorough content analyses in the future.

## Discussion

As many classification error occur because the tweets lie in the boundary between two classes, we note that a multi-label classification scheme might improve the performance.^47^ However, in the experiments conducted earlier in this project, we found the multi-label scheme only improved the classification performance by a small margin. We thus focused on single-label classification scheme in this work, leaving developing multi-label models in future work.

Besides multi-label classification models, the classification error might also be remedied by creating new categories for the tweets lying in the boundary of current categories. For example, we can further divide the political discussion (class *p*) into two categories, discussion of policy without personal experiences or experiences from friends or relatives and discussion with those experiences as supporting evidences. The classification performance can be further improved by including more user profile information. For example, we can include features such as if the account belongs to new agency or if the user has affiliation with academic organization or think tanks, which could improve the classification performance on class *n* or class *a*. As the two corpora have very different distributions, developing corpus specific classifier might also further improve the performance.

The content analysis, though now is limited to the high ranking TFIDF terms, could be further extended to include topic modeling to understand the recurring topics in the chatter,^48^ or sentimental analysis to understand the general sentiment toward Medicaid in general or the specific aspects of Medicaid.^8^ The manual analysis on selected samples can also deepen the understanding of these topics and potentially generate recommendation toward policy change. We also note that content analysis not only can help researchers further understand the Medicaid chatter, but also could improve the classification performance in reverse.

### Limitations

This work, however, is limited by the data, i.e. the tweets from Twitter users who choose express their opinions toward Medicaid publicly. The Twitter users are mostly millennials; the senior age group is underrepresented.^49^ Also, the data is from users who do not fear the retaliation against commenting on Medicaid publicly. Though it may not be an issue for many, the most vulnerable, the population who need Medicaid the most, might choose not to speak out and thus be left out by this work. We also note that our choice of categories and annotation guidelines are in no way perfect; we expect they will be modified and updated iteratively as we understand the chatter more.

### Conclusion

We have developed a social media mining system, involving NLP and machine learning, for continuously collecting and categorizing Twitter chatter about the Medicaid program. Our study demonstrates that it is possible to collect data about a large, complex health services and coverage program like Medicaid, using Twitter to obtain close-to-real-time knowledge about consumer perceptions and opinions. The automatic classification of streaming data is crucial, specifically for smaller classes, such as *consumer feedback*, for studying targeted topics.

Our analysis can inform public health researchers on how to use public discussion about health programs and services like Medicaid. Similarly, our system can be deployed by research groups or Medicaid agencies for continuous, on-going research on evolution of the public opinions on social media (e.g. the impact of certain policy changes or rulings). We also note that, though this work focuses on Medicaid, our methods and open-source code can readily be applied to other health services.

## Data Availability

Annotated data and methods will be available for future studies once the manuscript is published

## Acknowledgements

YY conducted and directed the machine learning experiments, evaluations and data analyses, with assistance from MAA, AS and WH. YY, AS, MAA and WH contributed to the data collection, annotation and analyses. JMZ and DG provided their expertise in preparing the annotation guidelines and categories, and helped formulate the overarching objectives of the project. AS, JMZ and DG provided supervision for various aspects of the study. YY drafted the manuscript and all authors contributed to the final manuscript. The authors thank the support from the Robert Wood Johnson Foundation.

## Abbreviations

API: Application Programming Interface
BERT: Bidirectional Encoder Representations from Transformers
BLSTM: Bi-directional Long Short-Term Memory
KNN: K-Nearest Neighbor
MA: Medicaid agency
MCO: Managed Care Organization
NB: Naïve Bayes
NLP: Natural Language Processing
NN: Shallow Neural Networks
RF: Random Forest
SVM: Support Vector Machine
TFIDF: Term-Frequency-Inverse-Document-Frequency
US: United States

