## Supplementary Materials for "Developing an automatic system for classifying chatter about health services from Twitter: A case study for Medicaid"

Jane M. Zhu, MD, MPP, MSHP^2^

David Grande, MD, MPA^3^

Abeed Sarker, PhD^1,4^

^1^Department of Biomedical Informatics, School of Medicine, Emory University, Atlanta, GA;

^2^Division of General Internal Medicine and Geriatrics, Oregon Health & Science University, Portland, OR;

^3^Division of General Internal Medicine, Perelman School of Medicine, University of Pennsylvania, Philadelphia, PA;

^4^­Department of Biomedical Engineering, Georgia Institute of Technology and Emory University, Atlanta, GA;

*Corresponding author

Table 1: List of keywords used for collecting Corpus 1

| Type of keywords | keywords |
| --- | --- |
| Medicaid and its common misspellings | medicaid  medicade  medcaid  medicad  mediaid  mediciad  medicaide |
| Branded Names of Medicaid Agencies or Managed Care Organizations | firstcare  excellusbcbs  mohealthnet  badgercare  wellsense  medquest  denalicare  forwardhealth  medi-cal  mainecare  tenncare  kancare  alamedaalliance  hapmichigan  ghcscw  mywellsense  cencalhealth  mclarenhlthplan  bhpartnershipma  vapremierhealth |

Table 2: List of handles used for collecting Corpus 2

| Medicaid Agency | AHCCCSgov, IAHealthLink, MDMedicaid, MassHealth, MSMedicaid, OhioMedicaid, oksoonercare, CCOOregon, scmedicaid, TennCareRep, DVHAVermont, WA_Health_Care, HUSKYHealthCT, GADCH, CHCPF |
| --- | --- |
| Managed Care Organizations | PSHP, mercycareaz, YamhillCCO, PassportHealthP, SFHealthPlan, AmidaCareNY, AetnaMedicaidNC, magnolia_health, HNEinc, AmeriHealthLA, NetworkHealthWI, UPMCHealthPlan, Medi_CalHCO, ExcellusBCBS, LACareHealth, QuartzBenefits, CoordinatedCare, Buckeye_Health, MVPHealthCare, PartnershipHP, hapmichigan, HPofSanJoaquin, CHCTexas, HSCHealthCare, AnthemBC_News, SunHealthFL, SilverSummitHP, FidelisCare, PriorityHealth, Trilliumchp, HorizonNJHealth, LAHealthConnect, PresHealth, BCBSNM, ECCooperative, HPPlans, RMHP, MHS_Wisconsin, NHPRIHealth, bhpartnershipma, HealthShareOR, VAPremierHealth, IlliniCare, GHCSCW, ucaremn, CFHealthPlans, iCareHealthPlan, _KFHC, MDwiseInc, HSCSN_Inc, CalOptima, SunflowerHPlan, HealthfirstNY, TRUSTEDHP, NextLevelIL, affinityplan, CencalHealth, AbetterGateway, HealthChoiceUT, AmbetterTX, MyWellSense, AlohaCareHawaii, YourCare_Health, CrystalRunHP, HealthChoiceAZ, firstcare, CoCoHealth, BCBSTX, MHSIndiana, UPHealthPlan, MetroPlusHealth, NHhealthy, CDPHP, AskHMSA, myCHPW, McLarenHlthPlan, CAHandW, IEHP_healthcare, BlueCrossMN, setonfamily, BMCHealthNet |

**Additional Notes about Data**

When collecting streaming data (Corpus-1), we limited the tweets to be English by specifying parameter “languages=['en'].” When collecting the tweets mentioning the handles belonging to MAs or MCOs (Corpus-2), we didn’t limit the tweets to be English. However, we found that the dataset consists of only about 0.2% of the non-English tweets, estimated via manually examining 1,000 tweets.

We have checked the overlap between the two corpora and found that there are 50 tweets present in both, which is about 0.01% of the data.

The 80% train /20% validation split was performed by selecting 80% of the data randomly as the training set and the rest as the validation set. We notice that there is higher composition of Academic tweets in the validation set and we believe that it have occurred by chance in the splitting process.

During the study, we have identified different types of noise in different stages of the study and thus applied different filtering when obtaining the data for 1^st^ and 2^nd^ round of annotation and the test datasets. We suspect that this is the reason that the Corpus-1 data annotated in the 1^st^ round of annotation and the test set of Corpus-1 have slightly different compositions. When generating the test set of Corpus-1, we first removed the data annotated in the 1^st^ round and then randomly draw from the remaining data. We also note that our method of removing noise (irrelevant or repeated contents) is not exhaustive; we identified noise by investigating erroneous high frequency/TFIDF terms or users with large volume of posts manually. Therefore, there may be still certain amount of noise in the corpora. One evidence is that there are tweets classified as class *Other*.

We found that some tweets in the training data (~250 in 2^nd^ round of annotation) contains unexpected special characters (such as â). Though we couldn’t explain their origin, we didn’t expect they would affect classification performance negatively and chose to keep those tweets in the training data. We found that about 10 re-tweets slipped into the training/validation sets. We removed those tweets after 2^nd^ round of annotation.

Table 3: The optimal performance of using SMOTE with SVM on the validation set

| **Precision (.XX)** | | | | | | **Recall (.XX)** | | | | | | **F1 (.XX)** | | | | | | **Accuracy (%)** |
| --- | --- | --- | --- | --- | --- | --- | --- | --- | --- | --- | --- | --- | --- | --- | --- | --- | --- | --- |
| **Academic** | **Consumer** | **Information** | **News** | **Political** | **Other** | **Academic** | **Consumer** | **Information** | **News** | **Political** | **Other** | **Academic** | **Consumer** | **Information** | **News** | **Political** | **Other** |  |
| 22 | 42 | 34 | 72 | 89 | 63 | 23 | 57 | 49 | 75 | 87 | 47 | 22 | 48 | 40 | 74 | 88 | 54 | 77.4 |

Table 4: The hyper-parameters for each classifier

| Classifier | Hyper-parameters |
| --- | --- |
| NN | 3 hidden layers with numbers of units (32, 16, 8), solver = ‘lbfgs’, , alpha=1e-5 |
| RF | n_estimators = 100 |
| SVM | C=1, class_weight={'a':8,'c':1.5,'i':2,'n':1,'p':1,'o':1}, kernel=’rbf’ |
| KNN | n_neighbors=7 |
| NB | default |
| BLSTM | Tokenization:  num_words: 100,000, maxlen: 128  Word Embeddings:  Twitter Glove, 200 dimensional vectors  Model:  an embedding layer [Output Shape: (None, 128, 200)] followed by a bidirectional LSTM layer [Output Shape: (None, 200)], followed by a dense layer [Output Shape: (None, 6)]  training:  loss function: ‘categorical_crossentropy’  optimizer: ‘adam’  dropout rate: 0.2  recurrent dropout: 0.2  metrics: accuracy  epochs: 40 |
| BERT | Tokenization:  max_seq_lenth: 128  Model:  RoBERTa-large  Training:  train_batch_size: 8  num_train_epochs: 3  learning_rate: 1e-5 |

**Table 5**: Classification performances of the classifiers on the validation set and the test sets of Corpus-1 & 2.

| **Classification Algorithm** | **Validation Set** | **Precision (.XX)** | | | | | | **Recall (.XX)** | | | | | |
| --- | --- | --- | --- | --- | --- | --- | --- | --- | --- | --- | --- | --- | --- |
|  |  | **Academic** | **Consumer** | **Information** | **News** | **Political** | **Other** | **Academic** | **Consumer** | **Information** | **News** | **Political** | **Other** |
|  | **NB** | 17 | 10 | 16 | 54 | 88 | 33 | 9 | 57 | 53 | 57 | 59 | 34 |
|  | **SVM** | 0 | 51 | 47 | 68 | 82 | 91 | 0 | 54 | 18 | 73 | 93 | 28 |
|  | **RF** | 50 | 54 | 50 | 73 | 81 | 86 | 3 | 35 | 18 | 75 | 94 | 33 |
|  | **KNN** | 50 | 22 | 15 | 49 | 89 | 16 | 3 | 27 | 8 | 63 | 51 | 66 |
|  | **NN** | 38 | 28 | 29 | 70 | 88 | 47 | 26 | 43 | 35 | 73 | 85 | 46 |
|  | **BLSTM** | 60 | 33 | 49 | 71 | 86 | 75 | 17 | 46 | 37 | 77 | 90 | 41 |
|  | **BERT** | 67 | 51 | 59 | 79 | 92 | 91 | 46 | 76 | 69 | 86 | 92 | 53 |
|  | **test set (Corpus-1)** | **Precision (.XX)** | | | | | | **Recall (.XX)** | | | | | |
|  |  | **Academic** | **Consumer** | **Information** | **News** | **Political** | **Other** | **Academic** | **Consumer** | **Information** | **News** | **Political** | **Other** |
|  | **NB** | 14 | 15 | 14 | 51 | 85 | 21 | 10 | 48 | 33 | 56 | 61 | 20 |
|  | **SVM** | 0 | 42 | 44 | 67 | 77 | 69 | 0 | 35 | 8 | 76 | 91 | 11 |
|  | **RF** | 0 | 36 | 67 | 74 | 76 | 75 | 0 | 17 | 12 | 76 | 94 | 14 |
|  | **KNN** | 0 | 21 | 21 | 43 | 89 | 16 | 0 | 20 | 12 | 51 | 53 | 66 |
|  | **NN** | 33 | 43 | 37 | 69 | 83 | 34 | 20 | 46 | 31 | 72 | 84 | 30 |
|  | **BLSTM** | 43 | 35 | 33 | 66 | 80 | 59 | 15 | 30 | 14 | 77 | 89 | 20 |
|  | **BERT** | 81 | 60 | 61 | 75 | 88 | 72 | 65 | 57 | 55 | 85 | 91 | 40 |
|  | **test set**  **(Corpus-2)** | **Precision (.XX)** | | | | | | **Recall (.XX)** | | | | | |
|  |  | **Academic** | **Consumer** | **Information** | **News** | **Political** | **Other** | **Academic** | **Consumer** | **Information** | **News** | **Political** | **Other** |
|  | **NB** | 0 | 82 | 20 | 7 | 2 | 47 | 0 | 64 | 68 | 24 | 17 | 13 |
|  | **SVM** | 0 | 66 | 50 | 14 | 4 | 60 | 0 | 89 | 1 | 38 | 67 | 11 |
|  | **RF** | 0 | 84 | 60 | 19 | 6 | 60 | 0 | 80 | 4 | 14 | 67 | 72 |
|  | **KNN** | 0 | 84 | 0 | 11 | 0 | 34 | 0 | 24 | 0 | 5 | 0 | 91 |
|  | **NN** | 0 | 87 | 47 | 21 | 3 | 66 | 0 | 72 | 35 | 29 | 50 | 66 |
|  | **LSTM** | 0 | 76 | 40 | 15 | 2 | 74 | 0 | 88 | 29 | 38 | 17 | 43 |
|  | **BERT** | 50 | 93 | 60 | 29 | 12 | 79 | 50 | 88 | 41 | 52 | 83 | 79 |

Table 6: The 10 highest-ranking TFIDF bigrams and trigrams in each class

| **Class** | **bigrams** | **trigrams** |
| --- | --- | --- |
| a | study finds  work requirements  study shows  states expanded  study says  health care  study suggests  low income  expansion states  affordable care | every state expanded  deaths every state  could averted deaths  heart related deaths  tied fewer heart  fewer heart related  expansion tied fewer  deaths study finds  study could averted  averted deaths every |
| c | health insurance  private insurance  food stamps  health care  social security  make much  full time  doesn cover  even though  nursing home | make much money  need transplant diabetes  knowing need transplant  healthcare knowing need  pocket drowning debt  diabetes leaving expansion  drowning debt please  leaving expansion medical  took healthcare knowing  open took healthcare |
| i | centers services  managed care  health care  long term  term care  health insurance  nursing home  critical carecoordination  carecoordination model  txmedicaidworks txlege | long term care  carecoordination model elements  critical carecoordination model  managed care members  model elements managed  elements managed care  assets devastating nursing  family assets devastating  devastating nursing home  protect family assets |
| n | work requirements  health care  trump administration  judge blocks  social security  federal judge  managed care  blocks work  kentucky arkansas  trump budget | judge blocks work  work requirements kentucky  requirements kentucky arkansas  social security budget  trump said wouldn  said wouldn social  blocks work requirements  security budget cuts  voter approved expansion  wouldn social security |
| p | social security  food stamps  health care  health insurance  cuts social  private insurance  trump budget  take away  pre existing  budget cuts | cuts social security  billion social security  pre existing conditions  cutting social security  welfare food stamps  trillion billion billion  billion billion social  social security budget  social security public  social security cuts |
